# A claims-based score for the prediction of bleeding in a contemporary cohort of patients receiving oral anticoagulation for venous thromboembolism

**DOI:** 10.1101/2021.02.01.21250924

**Authors:** Alvaro Alonso, Faye L. Norby, Richard F. MacLehose, Neil A. Zakai, Rob F. Walker, Terrence J. Adam, Pamela L. Lutsey

## Abstract

**Background:** Current scores for bleeding risk assessment in patients with venous thromboembolism (VTE) undergoing oral anticoagulation (OAC) have limited predictive capacity. We developed and internally validated a bleeding prediction model using healthcare claims data.

**Methods and Results:** We selected patients with incident VTE in the 2011-2017 MarketScan databases initiating OAC. Hospitalized bleeding events were identified using validated algorithms in the 180 days after VTE diagnosis. We evaluated demographic factors, comorbidities, and medication use prior to OAC initiation as potential predictors of bleeding using stepwise selection of variables in Cox models ran on 1000 bootstrap samples of the patient population. Variables included in >60% of all models were selected for the final analysis. We internally validated the model using bootstrapping and correcting for optimism. We included 165,434 VTE patients initiating OAC, of which 2,294 had a bleeding event. After undergoing the variable selection process, the final model included 20 terms (15 main effects and 5 interactions). The c-statistic for the final model was 0.68 (95% confidence interval [CI] 0.67-0.69). The internally validated c-statistic corrected for optimism was 0.68 (95%CI 0.67-0.69). For comparison, the c-statistic of the HAS-BLED score in this population was 0.62 (95%CI 0.61-0.63).

**Conclusion:** We have developed a novel model for bleeding prediction in VTE using large healthcare claims databases. Performance of the model was moderately good, highlighting the urgent need to identify better predictors of bleeding to inform treatment decisions.

## INTRODUCTION

One in twelve individuals will develop venous thromboembolism (VTE) during their lifetime.^1^ Oral anticoagulation (OAC) is the cornerstone of treatment for patients with VTE, with current guidelines recommending that most of these patients receive at least three to six months of anticoagulation after their diagnosis.^2^ Despite the potential risk of bleeding, the consequences of not treating acute VTE are severe enough that most individuals warrant anticoagulation for the primary treatment of VTE. Bleeding risk varies by choice of OAC with some of the newer oral agents having a lower major bleeding risk than warfarin.^3^ Bleeding risk factors may also differ by anticoagulant choice. Therefore, accurately characterizing individual patients’ bleeding risk is key to tailoring individualized treatment choices for the management of acute VTE.^2^ Over the years, several clinical prediction scores for major bleeding in patients with VTE have been developed to assist clinicians in this decision. These scores, however, were developed in cohorts with limited follow-up, did not compare bleeding risk across multiple oral anticoagulants, included small numbers of patients and bleeding events, and overall showed poor ability to discriminate risk.^4^ The current guideline from the American College of Chest Physicians for VTE treatment does not specifically recommend the use of any of these scores. Instead, the guideline categorizes patients according to the number of risk factors for bleeding as low risk (no risk factors), moderate risk (one risk factor), and high risk (two or more risk factors).^2^ Nonetheless, considerable variability in bleeding risk exists within each of these categories. Developing novel predictive models that quantify more accurately the risk of bleeding when receiving OAC is thus key to improve the care of people with acute VTE. To address this unmet need, we developed and internally validated a model for the prediction of bleeding in patients with VTE using a large healthcare claims database.

## METHODS

### Study population

This study was conducted within the IBM MarketScan® Commercial Claims and Encounters and Medicare Supplemental and Coordination of Benefits databases for the years 2011 through 2017. The MarketScan databases include individual-level HIPAA-compliant healthcare claims information from employers, health plans, hospitals, and Medicare programs from across the United States.^5^ Individual-level identifiers allow linkage across enrollment information and inpatient, outpatient, and pharmacy claims. The University of Minnesota Institutional Review Board deemed this research exempt from review, and waived the need to obtain informed consent.

This analysis included individuals 18 years of age and older with a diagnosis of VTE, at least one OAC prescription within one month after VTE, no use of OAC prior to VTE diagnosis, and ≥90 days of continuous enrollment prior to their first OAC prescription. We excluded dabigatran users due to small numbers (N = 1141); there were no users of edoxaban. Patient follow-up was censored at 180 days after VTE diagnosis. We defined VTE as having at least one inpatient claim or two outpatient claims 7 to 185 days apart including any International Classification of Diseases (ICD) code for VTE (Supplementary Table 1) in any position. A validation study using a similar definition of VTE reported a 91% positive predictive value for this algorithm.^6^

### Major bleeding events

The endpoint of interest was hospitalization for intracranial hemorrhage, gastrointestinal bleeding, or other major bleeding. Intracranial hemorrhage was defined as ICD-9-Clinical Modification (CM) codes 430.xx, 431.xx, or 432.xx or ICD-10-CM I60.xx, I61.xx, I62.xx as the primary discharge diagnosis in an inpatient claim, with a positive predictive value of this definition estimated to be >90%.^7^ Gastrointestinal bleeding and other major bleeding were defined using a previously described algorithm for identification of OAC-related bleeding that considers primary and secondary diagnosis in inpatient claims as well as the presence of transfusion codes.^8^ Positive predictive value of this algorithm is close to 90%.^8^

### Predictors

We identified potential predictors of bleeding from prior literature and existing risk scores. Predictors were defined according to validated algorithms when available using inpatient and outpatient ICD diagnosis codes, and pharmacy claims.^9, 10^ Specifically, we considered the following 24 predictors: age, sex, hypertension, diabetes, chronic kidney disease, myocardial infarction, heart failure, ischemic stroke, transient ischemic attack, peripheral artery disease, chronic obstructive pulmonary disease, liver disease, cancer, previous bleeding, anemia, excessive alcohol consumption, thrombocytopenia, and peptic ulcer disease, and the following medications: OAC type (warfarin, rivaroxaban, apixaban), antiplatelets, non-steroidal anti-inflammatory drugs, gastroprotective drugs (H2 receptor blockers, proton pump inhibitors, others), selective serotonin reuptake inhibitors, and cytochrome p450 3A4 inhibitors (atazanavir, clarithromycin, indinavir, itraconazole, ketoconazole, nefazodone, ritonavir, saquinavir, buprenorphine, telithromycin). We calculated the HAS-BLED (Hypertension, Abnormal renal/liver function, Stroke, Bleeding history or predisposition, Labile international normalized ratio, Elderly (>65 years), Drugs/alcohol concomitantly) score based on claims-derived diagnoses, with the exception of labile international normalized ratio due to unavailability of this information.^11^ Supplementary Table 2 provides a list of ICD-9-CM and ICD-10-CM codes used to define these covariates.

### Statistical analysis

We followed patients who initiated OAC after a VTE diagnosis from the time of OAC initiation to first occurrence of major bleeding hospitalization, day 180 post-VTE diagnosis, or December 31, 2017, whichever occurred earlier.

To select predictors of bleeding risk, we ran a Cox proportional hazards model including all the potential predictors listed above with stepwise backward selection of variables using p<0.05 as the inclusion threshold. This process was repeated in 1000 bootstrap samples of the study population and predictors included in more than 60% of the samples were selected for the final model.^12^ Once the initial list of predictors for the final models was selected via this process, we examined interactions between age, sex, OAC type and each one of the selected predictors. Individual interactions that were significant at p<0.05 were simultaneously added to the final model and those remaining statistically significant were kept. We evaluated the discriminatory value of the model by calculating the c-statistic and model calibration by comparing observed versus predicted probabilities by deciles of predicted risk. Model-based individual 180-day bleeding risk was calculated using the Breslow estimator, which is based on the empirical cumulative hazard function.^13^

Since we did not have access to an external data set, we performed an internal validation as recommended in existing guidelines for reporting of predictive models.^14^ Internal validation was done by creating 500 bootstrap samples of the study population and calculating the c-statistic in each sample using the model derived in the previous step. Because the model was derived and validated in the same dataset, we corrected the c-statistic for optimism.^15^

To facilitate comparison of the discriminative ability of the new model with that of a predictive model commonly used by clinicians, we calculated the c-statistic using the HAS-BLED score.

## RESULTS

The initial sample included 514,274 VTE patients older than 18 years of age. After restricting to OAC users, the sample was comprised of 401,013 patients. Requiring >90 days of enrollment prior to the first OAC prescription and excluding dabigatran users led to a final sample size of 165,434 VTE patients. Follow-up was censored at 180 days after VTE diagnosis, which was attained by 76% of patients. During a mean (standard deviation) follow-up time of 158 (46) days, we identified 2,294 bleeding events (3.2 events per 100 person-years). Figure 1 provides a flowchart of patient inclusion in the analysis.

**Figure 1.**
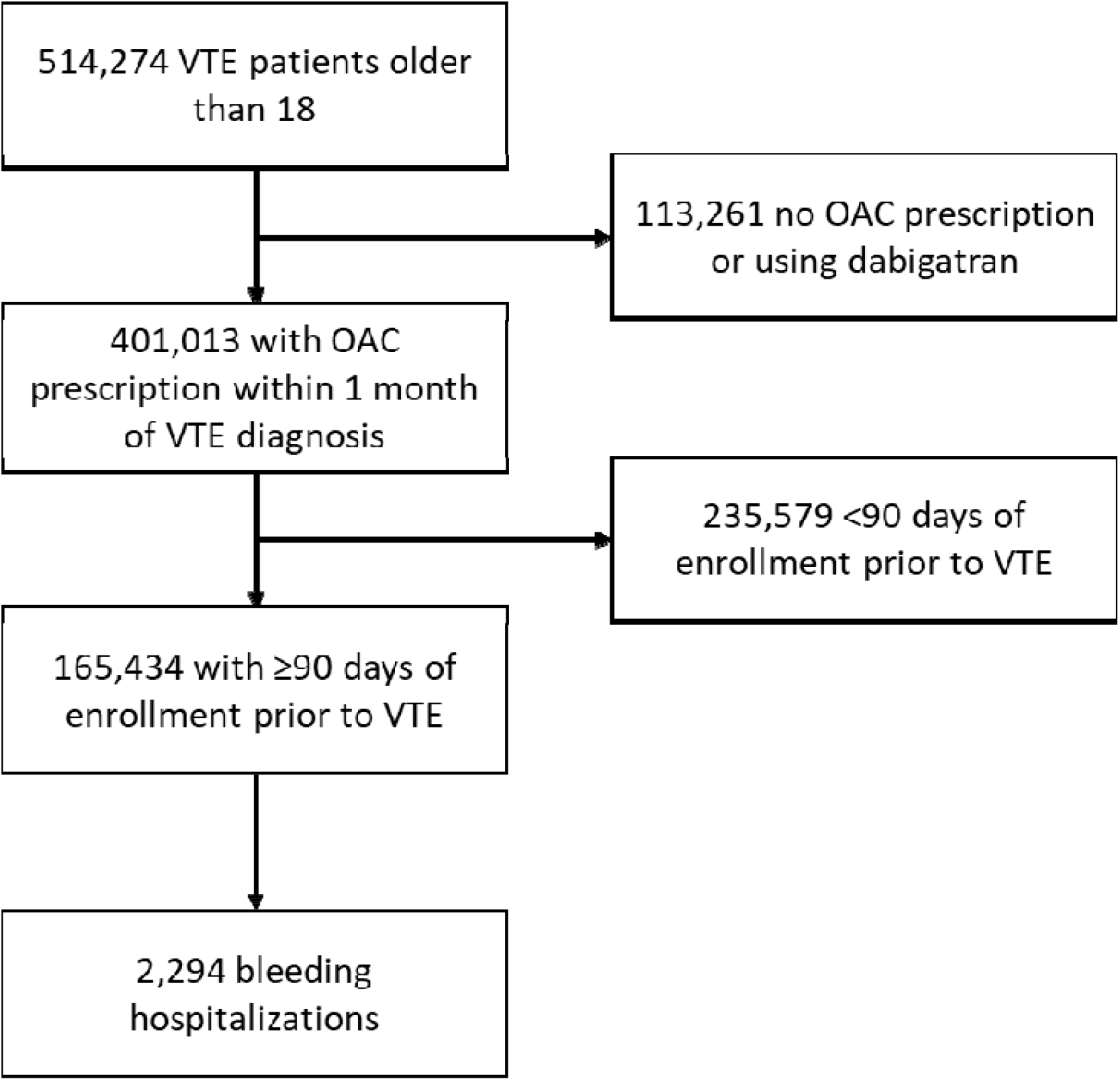
Flowchart of patient inclusion, MarketScan 2011-2017.

Table 1 shows descriptive characteristics of study patients overall and by type of OAC. Mean age (standard deviation) of patients was 58 (16) and 50% were female. The mean (standard deviation) HAS-BLED score was 1.7 (1.3). Patient characteristics across type of OAC were similar, except a slightly younger age and lower HAS-BLED score in rivaroxaban users than warfarin or apixaban users.

**Table 1.** Characteristics* of VTE patients by anticoagulant use, MarketScan 2011-2017

|  | Overall | Warfarin | Rivaroxaban | Apixaban |
| --- | --- | --- | --- | --- |
| N | 165,434 | 116,319 | 37,214 | 11,901 |
| Age, years | 58 ± 16 | 59 ± 16 | 56 ± 15 | 60 ± 16 |
| Female | 50 | 50 | 49 | 50 |
| Hypertension | 57 | 57 | 55 | 63 |
| Diabetes | 22 | 23 | 20 | 24 |
| Alcohol abuse | 0.9 | 0.7 | 1.2 | 2.2 |
| Myocardial infarction | 6.5 | 6.7 | 5.4 | 7.9 |
| Heart failure | 13 | 14 | 10 | 16 |
| Ischemic stroke/TIA | 11 | 12 | 9 | 8 |
| Renal Disease | 10 | 11 | 7.0 | 13 |
| Peripheral artery disease | 13 | 13 | 11 | 15 |
| Chronic pulmonary disease | 27 | 27 | 26 | 27 |
| Liver disease | 8.8 | 8.6 | 9.4 | 9.6 |
| Malignancy / Metastatic cancer | 18 | 18 | 16 | 17 |
| Anemia | 26 | 27 | 24 | 28 |
| Thrombocytopenia | 4.2 | 4.2 | 3.7 | 4.9 |
| Peptic ulcer disease | 0.7 | 0.7 | 0.7 | 0.7 |
| Other previous bleeding | 11 | 11 | 8.7 | 13 |
| HAS-BLED score | 1.7 ± 1.3 | 1.7 ± 1.3 | 1.6 ± 1.3 | 1.8 ± 1.3 |
| Warfarin | 70 | 100 | 0 | 0 |
| Rivaroxaban | 22 | 0 | 100 | 0 |
| Apixaban | 7.1 | 0 | 0 | 100 |
| Antiplatelets | 6.2 | 6.6 | 4.9 | 6.6 |
| NSAIDs | 35 | 33 | 41 | 34 |
| Gastroprotective drugs | 29 | 29 | 29 | 31 |
| SSRIs | 28 | 28 | 28 | 28 |
| Cyp3A4 inhibitors | 3.4 | 3.3 | 4.0 | 2.5 |
| * Values correspond to mean ± standard deviation or percentage. |  |  |  |  |

After running a stepwise Cox regression model in 1000 bootstrap samples, 15 variables were selected in >60% of the samples. Age, cancer history, anemia and type of OAC were selected in all predictive models. Other variables selected in >90% of the models included antiplatelets, liver disease, diabetes, previous bleeding and chronic pulmonary disease. The remaining selected variables were renal disease, alcohol abuse, female sex, prior ischemic stroke/TIA, and thrombocytopenia (Table 2).

**Table 2.** Predictors of bleeding considered in Cox regression models, MarketScan 2011-2017. Number of samples indicates the times that a variable was included in any of the 1000 bootstrap samples. Beta coefficient and hazard ratio (HR) and 95% confidence interval (CI) is for the final model including all covariates selected in >60% of the models.

| **Predictor** | **No. samples** | **Beta coefficient** | **HR (95%CI)** |
| --- | --- | --- | --- |
| **Age, per year** | 1000 | 0.011 | 1.01 (1.008, 1.014) |
| **Malignancy / Metastatic cancer** | 1000 | 0.355 | 1.43 (1.30, 1.57) |
| **Anemia** | 1000 | 0.500 | 1.65 (1.51, 1.81) |
| **Rivaroxaban (vs. warfarin)** | 1000 | -0.155 | 0.86 (0.77, 0.95) |
| **Apixaban (vs. warfarin)** | 1000 | -0.635 | 0.53 (0.43, 0.65) |
| **Antiplatelets** | 998 | 0.375 | 1.46 (1.27, 1.66) |
| **Liver disease** | 996 | 0.319 | 1.38 (1.22, 1.55) |
| **Diabetes** | 991 | 0.223 | 1.25 (1.14, 1.37) |
| **Other previous bleeding** | 986 | 0.265 | 1.30 (1.17, 1.46) |
| **Chronic pulmonary disease** | 930 | 0.182 | 1.20 (1.10, 1.31) |
| **Renal Disease** | 896 | 0.213 | 1.24 (1.11, 1.39) |
| **Alcohol abuse** | 857 | 0.547 | 1.73 (1.26, 2.36) |
| **Female** | 818 | 0.130 | 1.14 (1.05, 1.24) |
| **Ischemic stroke/TIA** | 740 | 0.163 | 1.18 (1.05, 1.32) |
| **Thrombocytopenia** | 607 | 0.194 | 1.21 (1.03, 1.43) |
| NSAIDs | 552 |  |  |
| Gastroprotective drugs | 520 |  |  |
| Heart failure | 462 |  |  |
| Peptic ulcer disease | 422 |  |  |
| SSRIs | 397 |  |  |
| Hypertension | 222 |  |  |
| Myocardial infarction | 139 |  |  |
| Peripheral artery disease | 88 |  |  |
| Cyp3A4 inhibitors | 42 |  |  |

Testing for interactions between age, sex, OAC class and the covariates selected in the final model identified 10 interactions with p<0.05 (Supplementary Table 3), most of them between age and comorbidities. After including these interactions in the final model, five of them remained significant. Table 3 shows the beta coefficients and p-values for all the significant predictors and their interactions in the final model. We have developed an Excel calculator that allows calculation of the predicted bleeding risk based on the patient characteristics (Supplementary Materials)

**Table 3.** Beta coefficients, standard errors, and p-values for bleeding predictors selected in final model, MarketScan 2011-2017.

| Predictor | Beta coefficient | SE | p-value |
| --- | --- | --- | --- |
| Age, per year | 0.021 | 0.002 | <0.001 |
| Female | 0.211 | 0.051 | <0.001 |
| Diabetes | 0.216 | 0.047 | <0.001 |
| Alcohol abuse | 0.528 | 0.160 | 0.001 |
| Ischemic stroke/TIA | 0.182 | 0.057 | 0.001 |
| Renal Disease | 0.233 | 0.058 | <0.001 |
| Chronic pulmonary disease | 0.184 | 0.045 | <0.001 |
| Liver disease | 0.294 | 0.062 | <0.001 |
| Malignancy / Metastatic cancer | 1.318 | 0.234 | <0.001 |
| Anemia | 1.269 | 0.185 | <0.001 |
| Thrombocytopenia | 0.180 | 0.083 | 0.03 |
| Other previous bleeding | 1.192 | 0.232 | <0.001 |
| Rivaroxaban (vs. warfarin) | -0.182 | 0.059 | 0.002 |
| Apixaban (vs. warfarin) | -0.763 | 0.126 | <0.001 |
| Antiplatelets | 0.379 | 0.068 | <0.001 |
| Age*cancer | -0.012 | 0.003 | <0.001 |
| Age*anemia | -0.012 | 0.003 | <0.001 |
| Age*previous bleed | -0.016 | 0.004 | <0.001 |
| Female sex*cancer | -0.347 | 0.093 | <0.001 |
| Rivaroxaban*previous bleed | 0.212 | 0.141 | 0.13 |
| Apixaban*previous bleed | 0.577 | 0.238 | 0.02 |

SE: Standard error. The 1-year risk of bleeding can be calculated as $1 - (0.98768)^{\text{Exp}[0.021 * (\text{Age} - 58.2) + 0.211 * (\text{Female sex} - 0.499) + 0.216 * (\text{Diabetes} - 0.221) + 0.528 * (\text{Alcohol abuse} - 0.009) + 0.182 * (\text{Ischemic stroke/TIA} - 0.111) + 0.233 * (\text{Renal disease} - 0.101) + 0.184 * (\text{Chronic pulmonary disease} - 0.266) + 0.294 * (\text{Liver disease} - 0.088) + 1.318 * (\text{Cancer} - 0.177) + 1.269 * (\text{Anemia} - 0.264) - 0.180 * (\text{Thrombocytopenia} - 0.041) + 1.192 * (\text{Other previous bleeding} - 0.108) - 0.182 * (\text{Rivaroxaban} - 0.225) - 0.763 * (\text{Apixaban} - 0.072) + 0.379 * (\text{Antiplatelets} - 0.062) - 0.012 * (\text{Age} * \text{Cancer} - 11.5) - 0.012 * (\text{Age} * \text{Anemia} - 16.3) - 0.016 * (\text{Age} * \text{Previous bleed} - 6.57) - 0.347 * (\text{Female sex} * \text{Cancer} - 0.088) + 0.212 * (\text{Rivaroxaban} * \text{Previous bleed} - 0.020) + 0.577 * (\text{Apixaban} * \text{Previous bleed} - 0.009)]}$

The c-statistic for the final model including main effects and interactions was 0.68 (95% confidence interval [CI] 0.67, 0.69). Calibration of the model, assessed by comparing observed and predicted probabilities across deciles of predicted probabilities was adequate (Figure 2). Patients in the top two deciles of predicted risk were at particularly high risk of bleeding (>2% over 180 days). Figure 3 shows the cumulative risk of bleeding by categories of predicted risk (low or <1%, moderate or 1-<2%, and high or ≥2%). Patients in the high-risk category accounted for 24% of the sample and 48% of all the bleeding events. Corresponding figures were 36% and 35% for the moderate-risk group, and 40% and 17% for the low-risk group. Correcting the c-statistic for optimism using 500 bootstrap samples resulted in essentially the same discrimination (c-statistic: 0.68, 95%CI 0.67, 0.69).

**Figure 2.**
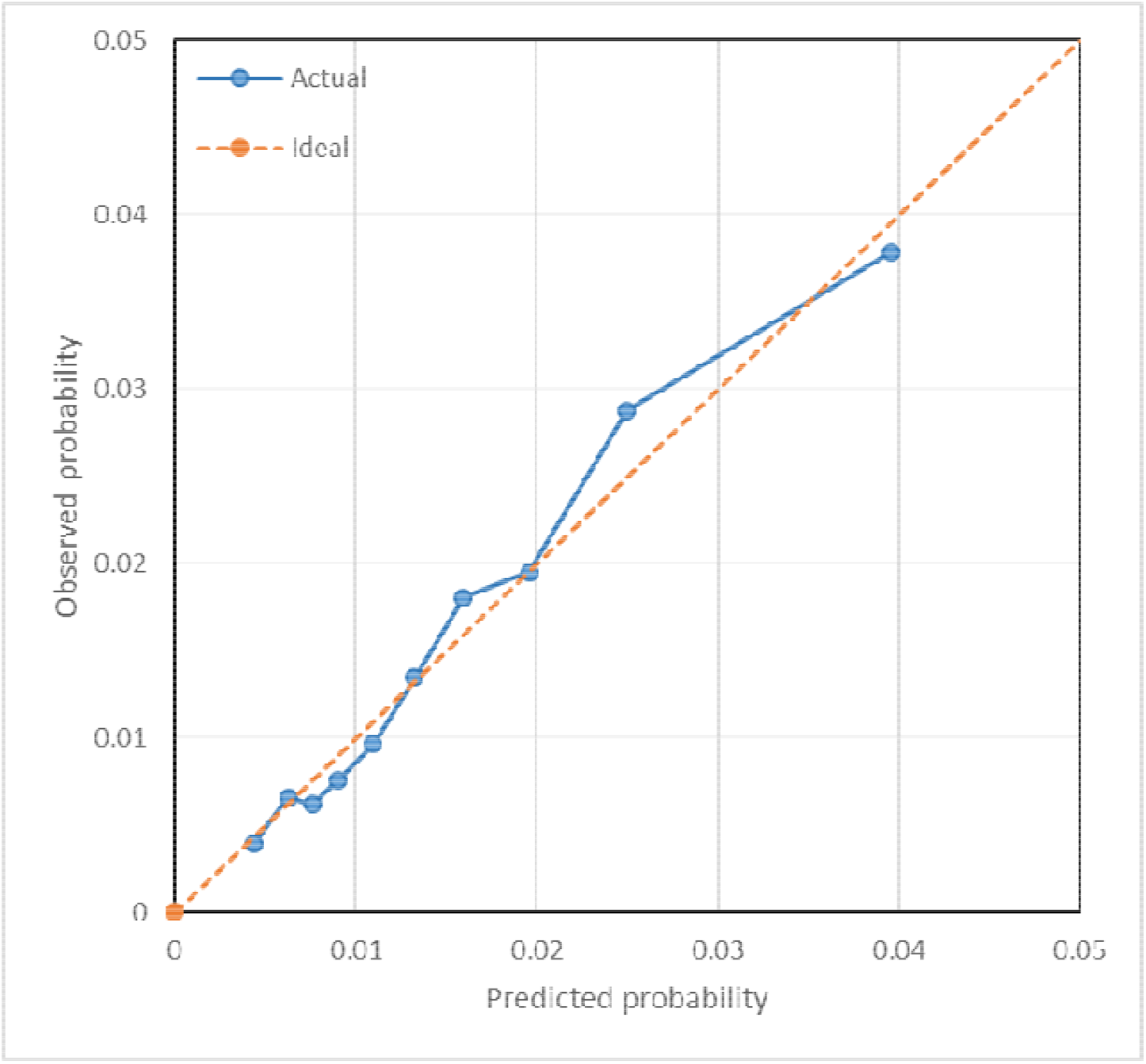
Calibration of predictive model, MarketScan 2011-2017. The plot shows the predicted versus observed probabilities by deciles of predicted risk (blue circles). Perfect calibration corresponds to the orange dashed line.

**Figure 3.**
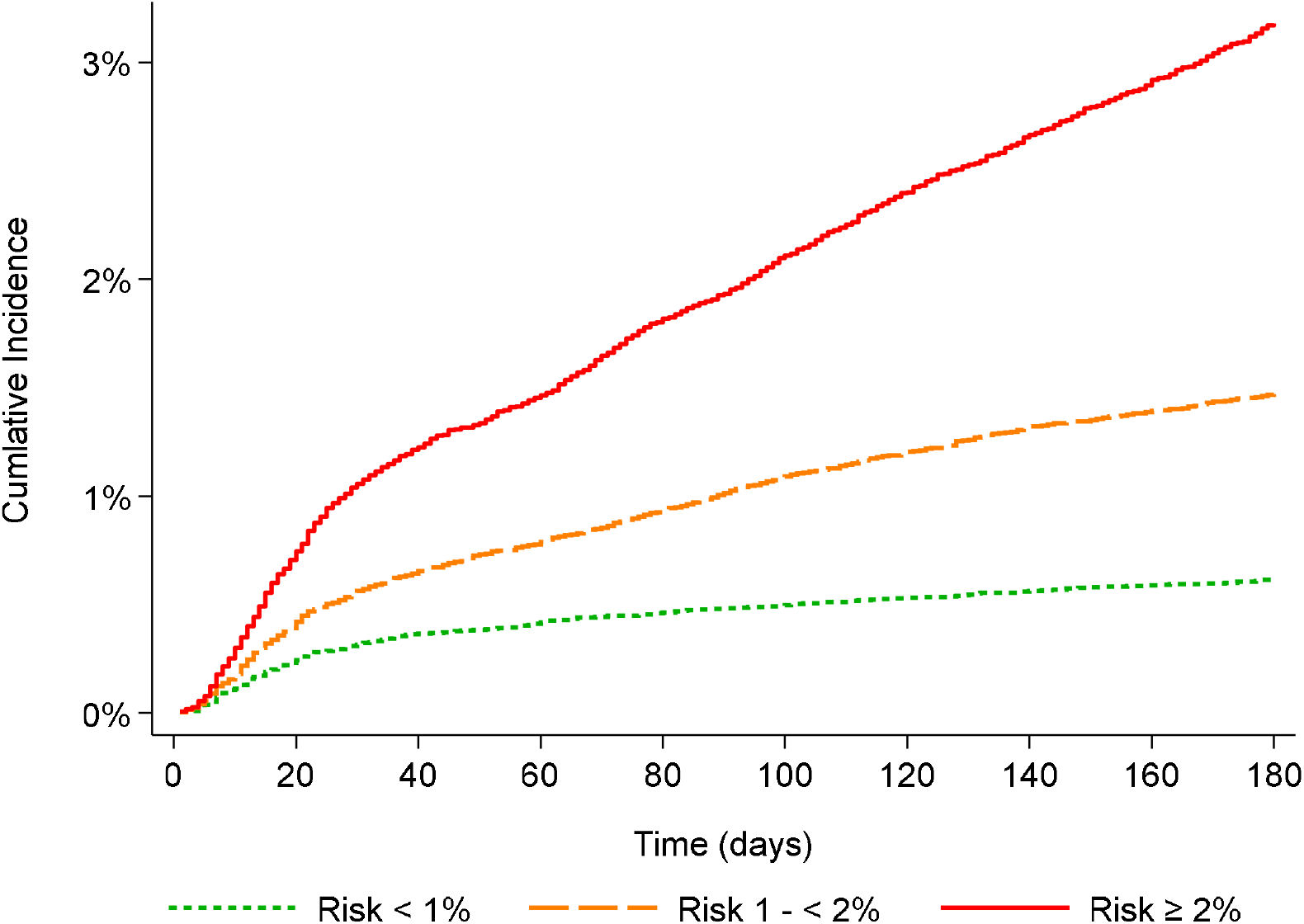
Cumulative incidence of hospitalized major bleeding by categories of 180-day predicted risk (<1%, 1-<2%, ≥2%), MarketScan 2011-2017

Discrimination of the model was similar in men and women, slightly better in younger patients, and in the direct OAC (DOAC) apixaban and rivaroxaban users compared to warfarin users (Table 4). Calibration was adequate across all subgroups of age category, sex and type of OAC.

**Table 4.** Discrimination and calibration of the prediction model overall and across subgroups.

| Group | C-statistic (95% CI) | Calibration $\chi^2$ (p-value) |
| --- | --- | --- |
| Full sample | 0.682 (0.671-0.692) | 27.6 (0.001) |
| Women | 0.666 (0.651, 0.681) | 9.1 (0.43) |
| Men | 0.699 (0.683, 0.714) | 14.4 (0.11) |
| Age $\leq 58$ | 0.688 (0.670, 0.707) | 16.3 (0.06) |
| Age $> 58$ | 0.652 (0.639, 0.666) | 16.7 (0.05) |
| Warfarin users | 0.669 (0.656, 0.681) | 15.9 (0.07) |
| Rivaroxaban users | 0.705 (0.679, 0.730) | 15.8 (0.07) |
| Apixaban users | 0.709 (0.657, 0.760) | 15.4 (0.08) |
| CI: 95% confidence interval |  |  |

The c-statistic for the HAS-BLED score (minus labile international normalized ratio) was 0.62 (95%CI 0.61, 0.63).

## DISCUSSION

We have developed and internally validated a model for the prediction of bleeding in patients with VTE based on information available in healthcare claims. The model identified a high-risk group that included approximately half of all the bleeding events in this patient cohort. In addition, the model performed similarly across different subgroups and had better discrimination than the established HAS-BLED score, in this dataset. The overall predictive ability of the model, however, was not exceptional, despite the inclusion of a large number of predictors.

Identifying patients with VTE at high risk of bleeding complications from OAC is an unmet clinical need. Existing predictive models and scores, such as the HAS-BLED score, have consistently shown mediocre performance when assessed by measures like the c-statistic.^16^ The model developed in this analysis performed slightly better than the HAS-BLED score, but not well enough to warrant extensive application. The limited ability of this newly developed algorithm and previous scores to predict major bleeding can be due, in part, to heterogeneity in the outcome, with different bleeding types having specific risk factors. Nonetheless, the information in the model has clinical relevance. First, it confirms that use of DOACs, particularly apixaban, instead of warfarin could result in overall lower bleeding risk in this patient group, as demonstrated in randomized trials and real-world effectiveness studies.^3, 17, 18^

Second, it identifies several comorbidities linked with increased bleeding risk. Whether better management of these comorbidities (e.g., anemia, diabetes, or alcohol abuse) results in lower bleeding risk merits further study. Third, our algorithm identified a significant interaction between prior bleeding and type of OAC, whereby the protective associations of apixaban and rivaroxaban are negated in patients with prior bleeding. This is a group underrepresented in clinical trials and, therefore, deserving of further study. Though a substudy of the ARISTOTLE trial did not find differences in the risk of bleeding of apixaban versus warfarin by prior bleeding history, this analysis was underpowered due to small number of patients with such history.^19^ Finally, being derived from claims data, this predictive model could be easily implemented and automatically calculated in electronic health record systems, making it easier to inform clinicians’ decisions.

In addition to the HAS-BLED score, there are several other scores for the prediction of bleeding in VTE patients receiving OACs.^4^ These scores generally include few variables and are weighted using a point system to facilitate clinical application. Our model, in contrast, includes a larger number of predictors than other scores and does not assign points to each variable. Rather, it uses precise information from the model coefficients to calculate risk in a predictive equation. This approach has the advantage of not discarding information and providing more accurate predictions. We have successfully used this approach in the past to develop models for prediction of bleeding and stroke in patients with atrial fibrillation.^10, 20^

The present analysis has some notable strengths. The model has been developed in a large population including a sizable number of bleeding events, one order of magnitude larger than many other published scores. It also takes advantage of the extensive information on comorbidities and medication use available through claims data. Finally, it incorporates DOACs as predictors, making it more applicable in the contemporary clinical setting. However, this analysis also has weaknesses, most importantly the lack of an external validation sample and the limitations in the validity of claims diagnoses to define bleeding and comorbidities, despite the use of validated algorithms. Comparisons with the HAS-BLED score are likely biased in favor of our algorithm since we developed and tested the algorithm in the same dataset and HAS-BLED was developed in a separate dataset. Other shortcomings are the inadequate sample size to properly evaluate model performance in subgroups, unavailability of information on laboratory values and clinical measurements, absence of information on over-the-counter drug use, and the potential lack of generalizability to uninsured individuals.

In conclusion, we developed a novel model to predict bleeding in VTE patients receiving OAC using healthcare claims information. This model predicted slightly better than a modified HAS-BLED score, but overall predictive performance was wanting. Additional work is needed to evaluate whether additional types of information, such as biomarkers, genetic factors, or drug therapy problem data, could improve predictive ability of this or other models. Also, future work should combine prediction of VTE recurrence and bleeding to facilitate decisions to clinicians and patients.

## Supporting information

Bleeding risk calculator

## Data Availability

Data used in this manuscript are available from IBM Watson Health, owner of the MarketScan database.

## REFERENCES

1. Bell EJ, Lutsey PL, Basu S, Cushman M, Heckbert SR, Lloyd-Jones DM, Folsom AR. Lifetime risk of venous thromboembolism in two cohort studies. Am J Med. 2016;129:339.e319-e326.

2. Kearon C, Akl EA, Ornelas J, Blaivas A, Jimenez D, Bounameaux H, Huisman M, King CS, Morris TA, Sood N, Stevens SM, Vintch JRE, Wells P, Woller SC, Moores L. Antithrombotic therapy for VTE disease: CHEST Guideline and Expert Panel Report. Chest. 2016;149:315–352.

3. Lutsey PL, Zakai NA, MacLehose RF, Norby FL, Walker RF, Roetker NS, Adam TJ, Alonso A. Risk of hospitalised bleeding in comparisons of oral anticoagulant options for the primary treatment of venous thromboembolism. Br J Haematol. 2019;185:903–911.

4. van Es N, Wells PS, Carrier M. Bleeding risk in patients with unprovoked venous thromboembolism: A critical appraisal of clinical prediction scores. Thromb Res. 2017;152:52–60.

5. IBM Watson Health. IBM MarketScan Research Databases for Health Services Research--White Paper. Somers, NY 2018.

6. Sanfilippo KM, Wang T-F, Gage BF, Liu W, Carson KR. Improving accuracy of international classification of diseases codes for venous thromboembolism in administrative data. Thromb Res. 2015;135:616–620.

7. Andrade SE, Harrold LR, Tjia J, Cutrona SL, Saczynski JS, Dodd KS, Goldberg RJ, Gurwitz JH. A systematic review of validated methods for identifying cerebrovascular accident or transient ischemic attack using administrative data. Pharmacoepidemiol Drug Saf. 2012;21 Supplement 1:100–128.

8. Cunningham A, Stein CM, Chung CP, Daugherty JR, Smalley WE, Ray WA. An automated database case definition for serious bleeding related to oral anticoagulant use. Pharmacoepidemiol Drug Saf. 2011;20:560–566.

9. Quan H, Sundararajan V, Halfon P, Fong A, Burnand B, Luthi JC, Saunders LD, Beck CA, Feasby TE, Ghali WA. Coding algorithms for defining comorbidities in ICD-9-CM and ICD-10 administrative data. Medical Care. 2005;43:1130–1139.

10. Claxton JNS, MacLehose RF, Lutsey PL, Norby FL, Chen LY, O’Neal WT, Chamberlain AM, Bengtson LGS, Alonso A. A new model to predict major bleeding in patients with atrial fibrillation using warfarin or direct oral anticoagulants. PLoS One. 2018;13:e0203599.

11. Pisters R, Lane DA, Nieuwlaat R, de Vos CB, Crijns HJGM, Lip GYH. A novel user-friendly score (HAS-BLED) to assess 1-year risk of major bleeding in patients with atrial fibrillation: The Euro Heart Survey. Chest. 2010;138:1093–1100.

12. Austin PC, Tu JV. Bootstrap methods for developing predictive models. Am Stat. 2004;58:131–137.

13. Breslow NE. Discussion of Professor Cox’s paper. J Royal Stat Soc B. 1972;34:216–217.

14. Collins GS, Reitsma JB, Altman DG, Moons KGM. Transparent reporting of a multivariable prediction model for individual prognosis or diagnosis (TRIPOD): the TRIPOD statement. BMJ. 2015;350:g7594.

15. Harrell FE, J., Lee KL, Mark DB. Multivariable prognostic models: issues in developing models, evaluating assumptions and adequacy, and measuring and reducing errors. Stat Med. 1996;15:361–387.

16. Vedovati MC, Mancuso A, Pierpaoli L, Paliani U, Conti S, Ascani A, Galeotti G, Di Filippo F, Caponi C, Agnelli G, Becattini C. Prediction of major bleeding in patients receiving DOACs for venous thromboembolism: A prospective cohort study. Int J Cardiol. 2020;301:167–172.

17. The EINSTEIN-PE Investigators. Oral rivaroxaban for the treatment of symptomatic pulmonary embolism. N Engl J Med. 2012;366:1287–1297.

18. Agnelli G, Buller HR, Cohen A, Curto M, Gallus AS, Johnson M, Masiukiewicz U, Pak R, Thompson J, Raskob GE, Weitz JI. Oral apixaban for the treatment of acute venous thromboembolism. N Engl J Med. 2013;369:799–808.

19. Garcia DA, Fisher DA, Mulder H, Wruck L, De Caterina R, Halvorsen S, Granger CB, Held C, Wallentin L, Alexander JH, Lopes RD. Gastrointestinal bleeding in patients with atrial fibrillation treated with Apixaban or warfarin: Insights from the Apixaban for Reduction in Stroke and Other Thromboembolic Events in Atrial Fibrillation (ARISTOTLE) trial. Am Heart J. 2020;221:1–8.

20. Claxton JS, MacLehose RF, Lutsey PL, Norby FL, Chen LY, O’Neal WT, Chamberlain AM, Bengtson LGS, Alonso A. A new model to predict ischemic stroke in patients with atrial fibrillation using warfarin or direct oral anticoagulants. Heart Rhythm. 2019; (in press).

