## Supplementary Tables for "A claims-based score for the prediction of bleeding in a contemporary cohort of patients receiving oral anticoagulation for venous thromboembolism"

**SUPPLEMENTARY MATERIALS**

Supplementary Table 1. International Classification of Disease (ICD) Clinical Modification (CM) diagnosis codes for venous thromboembolism (VTE).

| Version | VTE codes |
| --- | --- |
| ICD-9-CM | 415.1x, 451.1x, 453.2, 453.4x, 453.82, 453.83, 453.84, 453.85, 453.86,  453.87, 453.89, 453.9 |
| ICD-10-CM | I26.0x, I26.9x, I80.1x, I80.20x, I82.210, I80.22x, I80.23x, I80.29x, I82.40x, I82.41x, I82.42x, I82.43x, I82.44x, I82.49x, I82.4Yx, I82.4Zx, I82.60x, I82.62x, I82.890, I82.A1x, I82.B1x, I82.C1x |

Supplementary Table 2. International Classification of Disease (ICD) Clinical Modification (CM) diagnosis codes for comorbidities considered as potential predictors.

| **Condition** | **ICD-9-CM** | **ICD-10-CM** |
| --- | --- | --- |
| Alcohol abuse | 265.2, 291.1, 291.2, 291.3, 291.5, 291.6, 291.7, 291.8, 291.9, 303.0, 303.9, 305.0, 357.5, 425.5, 535.3, 571.0, 571.1, 571.2, 571.3, 980, V11.3 | F10, E52, G62.1, I42.6, K29.2, K70.0, K70.3, K70.9, T51.x, Z50.2, Z71.4, Z72.1 |
| Anemia | 280.x-284.x, 285.1, 285.2, 285.3, 285.8, 285.9 | D50.x-D53.x, D55.x-D64.x |
| Chronic pulmonary disease | 416.8, 416.9, 490.x-505.x, 506.4, 508.1, 508.8 | I27.8, I27.9, J40.x-J47.x, J60.x-J67.x, J68.4, J70.1, J70.3 |
| Diabetes | 250.x | E10.0-E10.9, E11.0-E11.9, E12.0-E12.9, E13.0-E13.9, E14.0-E14.9 |
| Heart failure | 398.91, 402.01, 402.11, 402.91, 404.01, 404.03, 404.11, 404.13, 404.91, 404.93, 425.4-425.9, 428.x | I09.9, I11.0, I13.0, I13.2, I25.5, I42.0, I42.5-I42.9, I43.x, I50.x, P29.0 |
| Hypertension | 401.x, 402.x, 403.x, 404.x, 405.x | I10.x, I11.x-I13.x, I15.x |
| Ischemic stroke / TIA | 362.34, 430.x-438.x | G45.x, G46.x, H34.0, I60.x-I69.x |
| Liver disease | 070.22, 070.23, 070.32, 070.33, 070.44, 070.54, 070.6, 070.9, 456.0, 456.1, 456.2, 570, 571, 572.2, 572.3, 572.4, 572.5, 572.6, 572.7, 572.8, 573.3, 573.4, 573.8, 573.9, V42.7 | B18.x, K70.0-K70.3, K70.9, K71.3-K71.5, K71.7, K73.x, K74.x, K76.0, K76.2-K76.4, K76.8, K76.9, Z94.4, I85.0, I85.9, I86.4, I98.2, K70.4, K71.1, K72.1, K72.9, K76.5, K76.6, K76.7 |
| Malignancy / metastatic cancer | 140.x-172.x, 174.x-195.8, 196.x-199.x, 200.x-208.x, 238.6 | C00.x-C26.x, C30.x-C34.x, C37.x-C41.x, C43.x, C45.x-C58.x, C60.x-C76.x, C77.x-C80.x, C81.x-C85.x, C88.x, C90.x-C97.x |
| Myocardial infarction | 410.x, 412.x | I21.x, I22.x, I25.2 |
| Peptic ulcer disease | 533.x | K27.0-K27.7, K27.9 |
| Peripheral artery disease | 093.0, 437.3, 440.x, 441.x, 443.1-443.9, 47.1, 557.1, 557.9, V43.4 | I70.x, I71.x, I73.1, I73.8, I73.9, I77.1, I79.0, I79.2, K55.1, K55.8, K55.9, Z95.8, Z95.9 |
| Renal disease | 403.01, 403.11, 403.91, 404.02, 404.03, 404.12, 404.13, 404.92, 404.93, 582, 583.0, 583.1, 583.2, 583.3, 583.4, 583.5, 583.6, 583.7, 585, 586, 588.0, V42.0, V45.1, V56 | I12.0, I13.1, N03.2-N03.7, N05.2-N05.7, N18.x, N19.x, N25.0, Z49.0-Z49.2, Z94.0, Z99.2 |
| Thrombocytopenia | 287.1, 287.3, 287.4, 287.5 | D69.3, D69.6 |

Supplementary Table 3. Significant interactions identified in the development of the prediction model. All interactions of age, sex, and OAC type with the variables included in the model were tested. Only those with p-value <0.05 were retained for further testing.

| Interactions | P for interaction |
| --- | --- |
| Age*ischemic stroke | 0.002 |
| Age*renal disease | 0.001 |
| Age*liver disease | 0.002 |
| Age*cancer | <0.0001 |
| Age*anemia | <0.001 |
| Age*thrombocytopenia | 0.04 |
| Age*previous bleed | <0.001 |
| Sex*liver disease | 0.04 |
| Sex*cancer | <0.001 |
| OAC class*previous bleed | 0.03 |
